# Circadian meal timing is heritable and associated with insulin sensitivity

**DOI:** 10.1101/2024.09.04.24312795

**Authors:** Janna Vahlhaus, Beeke Peters, Silke Hornemann, Anne-Cathrin Ost, Michael Kruse, Andreas Busjahn, Andreas F.H. Pfeiffer, Olga Pivovarova-Ramich

**Affiliations:** Department of Molecular Metabolism and Precision Nutrition, German Institute of Human Nutrition Potsdam-Rehbruecke, Nuthetal, Germany; University of Lübeck, Lübeck, Germany; German Center for Diabetes Research (DZD), München-Neuherberg, Germany; Department of Clinical Nutrition, German Institute of Human Nutrition Potsdam-Rehbruecke, Nuthetal, Germany; Charité – Universitätsmedizin Berlin, Corporate Member of Freie Universität Berlin and Humboldt-Universität zu Berlin, Department of Endocrinology, Diabetes and Nutrition, Campus Benjamin Franklin, Berlin, Germany; HealthTwiSt GmbH, Berlin, Germany; Charité – Universitätsmedizin Berlin, Corporate Member of Freie Universität Berlin and Humboldt-Universität zu Berlin, Department of Endocrinology and Metabolism, Berlin, Germany

**Keywords:** meal timing, circadian clocks, glucose metabolism, diabetes, chronotype, heritability, twins

## Abstract

**Background:** Although the contribution of the circadian clock to metabolic regulation is widely recognized, the role of meal timing in glucose metabolism and diabetes risk remains insufficiently studied. This study aimed (i) to investigate the link between individual circadian meal timing pattern and glucose homeostasis and (ii) to explore the contribution of genetic and environmental factors to meal timing parameters.

**Methods:** In the German NUtriGenomic Analysis in Twins (NUGAT) cohort, which includes 92 adult twins, glucose metabolism parameters were assessed using fasting samples and the oral glucose tolerance test (OGTT). Parameters of meal timing pattern (meal timing itself, daily calorie distribution, and meal number) were extracted from five-day food records. Circadian eating timing was determined relative to the individual’s chronotype (MSFsc) assessed by the Munich chronotype questionnaire. The heritability of meal timing components was estimated using the ACE model.

**Results:** Multiple meal timing components showed associations with glucose metabolism parameters. Most associations were found for the calorie midpoint defined as the time point at which 50% of daily calories were consumed. Indices of insulin sensitivity, ISI Stumvoll (β = 0.334, p = 2.9 x 10^-4^) and HOMA-IR (β = -0.276, p = 0.007), as well as fasting insulin levels were significantly associated with the circadian caloric midpoint even after the model adjustment for gender, age, energy intake, and sleep duration. BMI and waist circumference also demonstrated robust associations with circadian caloric midpoint. High or moderate heritability was shown for all meal timing components. Meal timing pattern was also strongly related to individual sleep timing and chronotype, both of which also showed a marked genetic impact.

**Conclusion:** Circadian meal timing is associated with insulin sensitivity and shows significant genetic influences, sharing a common genetic architecture with sleep behaviour. Shifting the main calorie intake to earlier circadian time might protect against diabetes, although this could be challenging due to the high heritability of meal timing components.

**Graphical abstract:** 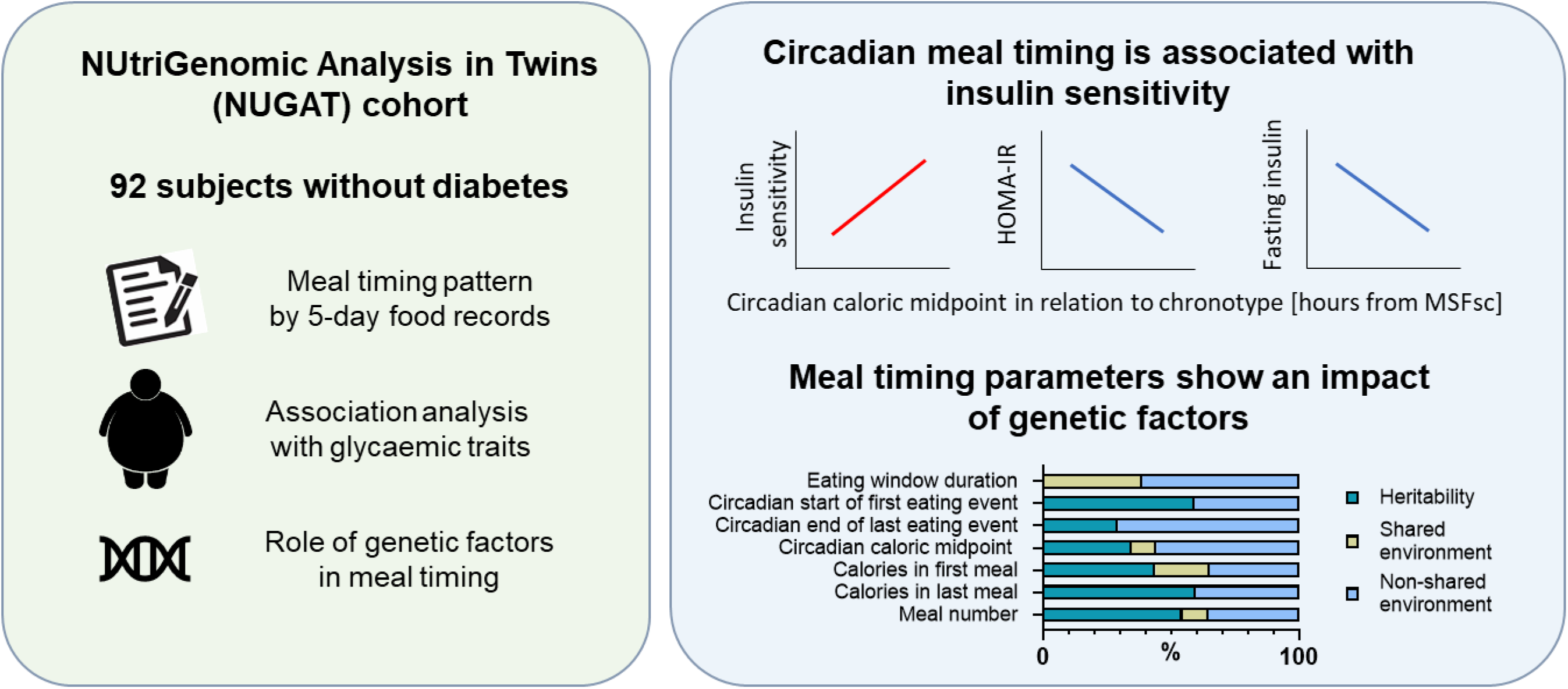

**Highlights:**

- Circadian caloric midpoint shows a robust association with insulin sensitivity
- It remains significant after the adjustment for energy intake and other cofounders
- Meal timing, daily calorie distribution, and meal number show a high or moderate heritability
- Meal timing strongly relates to the sleeping behaviour and chronotype

## 1. Introduction

The circadian system is a hierarchical 24-hour timing system in mammals, regulating corresponding daily rhythms of behaviour and metabolism through the central clock in the hypothalamus and peripheral clocks in different tissues and organs [1, 2]. Therefore, our body reacts differently to the same food consumed at different times of the day, showing diurnal variation in glucose tolerance, postprandial hormone secretion, thermogenesis, metabolite levels, and other metabolic processes [3, 4]. Further, food intake is one of most important cues (zeitgebers) entraining circadian clocks to synchronize them with the environment, especially affecting the clocks of the liver, gut, pancreas and adipose tissue [3, 5]. The desynchronization of meal timing with the external light-dark cycle (e.g. during night shift work) induces internal clock desynchronization and adverse metabolic events [6]. These associations between meal timing and metabolism have been confirmed by epidemiological and interventional studies, showing the relation of late and night eating to an increased risk of obesity, cardiovascular diseases, and related cardiometabolic traits such as higher BMI and body fat [7–12]. Nevertheless, how meal timing contributes to the parameters of glucose metabolism and generally to the diabetes risk remains poorly studied. Further, except of few studies [13], the impact of various components of the meal timing pattern, including timing, frequency, and diurnal calorie distribution, is still not investigated in detail.

Mechanisms determining meal timing pattern also still need to be clarified as they involve a complex interaction of cultural and environmental factors, behavioural and personal preferences (e.g., chronotype) as well as physiological factors [14]. To develop effective dietary interventions, it is of great interest to differentiate non-modifiable factors such as the heritability of meal timing [15]. To our knowledge, they have only been analysed in two studies, showed marked heritability for the timing of breakfast, lunch, and dinner [16, 17].

Therefore, the primary aim of this study was to assess the link between components of the meal timing pattern and glucose metabolism parameters. The second aim of this study was to explore the contribution of genetic and environmental factors to the individual’s meal timing pattern. For this, we used a NUtriGenomic Analysis in Twins (NUGAT) cohort combining a detailed characterization of the meal timing pattern, glucose homeostasis, and circadian phenotyping.

## 2. Materials and Methods

### 2.1. Study participants and study protocol

The NUGAT study was conducted between September 2009 and January 2010 at the German Institute of Human Nutrition Potsdam-Rehbruecke in accordance with the principles of the Declaration of Helsinki from 1975, as revised in 2000. The study protocol was approved by the ethics committee of the Charité-Universitätsmedizin Berlin and registered at ClinicalTrials.gov (unique identifier: NCT01631123). All participants were informed about the aim, content, and risks of the study and gave written consent. Twin pairs were recruited either from a twin register (HealthTwiSt, Berlin, Germany) or through public advertisements. Inclusion criteria were age 18 to 70 years and BMI between 18 and 30 kg/m², with BMI difference <3 kg/m^2^ between twins. Exclusion criteria included diabetes mellitus, high-grade anemia, renal failure, moderate to severe heart diseases, angina pectoris, or stroke in the last 6 months, food allergy, eating disorders, body weight change ≥3 kg within 3 months prior to the study, pregnancy and breastfeeding, and using of drugs influencing metabolic homeostasis as described in details elsewhere [18, 19].

The 112 subjects were initially screened to determine their eligibility, and 92 participants (46 twin pairs) were enrolled into the study (**Figure S1**). Zygosity was confirmed by DNA testing. Participants underwent a detailed metabolic phenotyping including physical examination, medical history, anthropometric measurements, and a standardized 3-hour, 75 g oral glucose tolerance test (OGTT). After the visit, all subjects completed handwritten dietary records, noting the start and end of each eating event, the amount and kind of consumed food, for five consecutive days including three working days and two free days to encompass their dietary habits. They did not receive any dietary recommendation or restrictions before completing the food records.

### 2.2. Anthropometric measurements

Body weight was measured with a digital scale and body height was measured with a stadiometer. Waist and hip circumferences were measured using a metric tape. Body composition was analysed via dual energy x-ray absorptiometry.

### 2.3. Blood sample analyses and assessment of glycaemic indices

Blood samples were drawn from the forearm vein after an overnight fast (>10 h since last food intake) and during the OGTT 30, 60, 90, 120 and 180 min after glucose intake. Routine serum parameters and plasma glucose were determined using an automated analyzer (ABX Pentra 4000; ABX, Montpellier, France). Serum insulin concentrations were analysed by ELISA (Mercodia, Uppsala, Sweden). Based on the glucose and insulin values, homeostasis model assessment of insulin resistance (HOMA-IR) was calculated as (Gluc_0_ (mmol/l) x Ins_0_ (µU/ml)) / 22.5. The insulinogenic index was calculated as Ins_30-0_ (µU/ml) /Gluc_30-0_ (mg/dl) [20]. The disposition index, which reflects the relation between β-cell secretion capacity and insulin sensitivity, was assessed as described [21]. The OGTT-derived Stumvoll index of insulin sensitivity (ISI Stumvoll), which shows one of the highest correlation with clamp-derived values, was calculated as 0.226 – 0.0032 x BMI – 0.0000645 x Ins_120_ (µU/ml) – 0.00375 x Gluc_90_ (mmol/l) [20]. Additionally, a Matsuda index of insulin sensitivity (ISI Matsuda) was calculated as described [22].The area under the curve (AUC) for glucose and insulin were assessed by trapezoidal method.

### 2.4. Analysis of meal timing pattern

Digitalization of food records was performed using the PRODI software version 4.5 (NutriScience GmbH, Hofstetten-Flüh, Switzerland) by the experienced nutritionist. Two food records contained no information on the meal timing, and one subject was a shift worker; therefore 89 records were included in the final analysis (**Figure S1**). The digital versions were used to assess total energy intake (in kcal) and macronutrient composition (in EN%) during the day and during each meal as well as clock timing of eating events. Eating window duration was calculated as time difference from the start of the first eating event to the end of the last eating event, and the caloric midpoint of calorie intake was assessed as time when the 50% of total calorie intake was reached [23]. Circadian timing of the first and last eating event and of the caloric midpoint was determined as a time interval to the midpoint of sleep corrected for sleep-debt accumulated over the week (MSFsc) (**Figure 1C**) which is a marker of individual circadian phase (chronotype) as described below (p.2.5).

**Figure 1.**
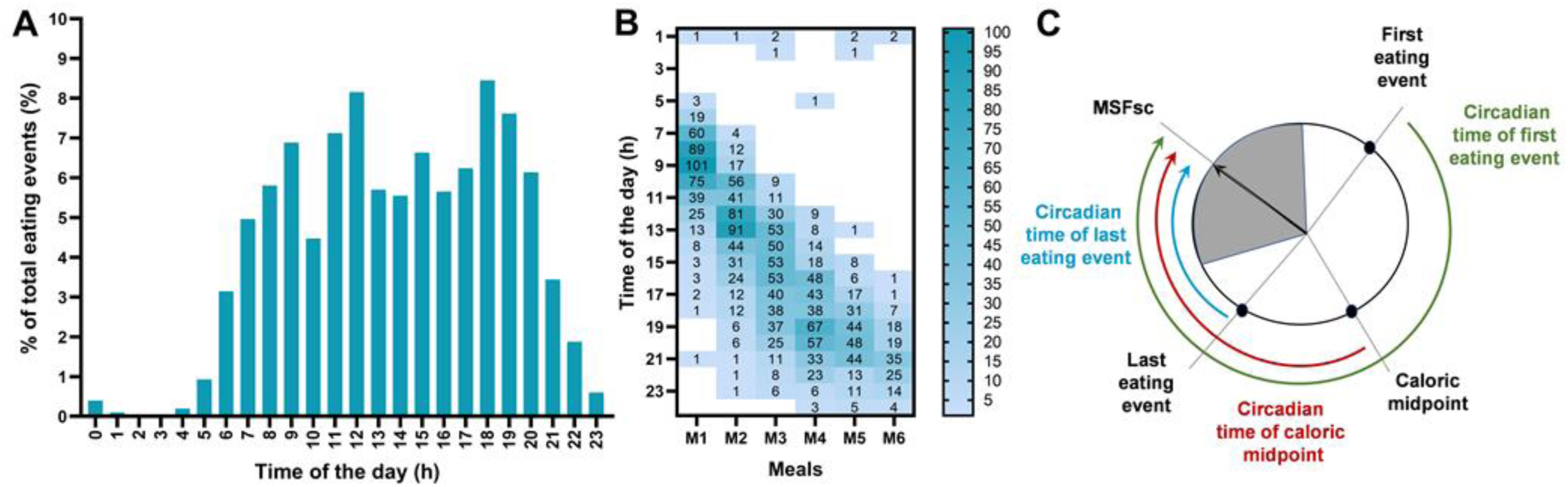
Daily distribution of eating events. Distribution of eating events is shown as percentage of all eating events (**A**) and in total number eating events classified as meals 1-6 (M1-6) (**B**). Each number at the time axis represents an hour starting from 0 (24:00 – 1:00) and ending with 23 (23:00 – 24:00). Some persons showed an untypical eating habit consuming only one to two meals in the evenings. (**C**) Calculation of circadian timing of the first and last eating events and of the caloric midpoint related to midpoint of sleep corrected for sleep-debt accumulated over the week (MSFsc). The grey shaded area denotes habitual sleep timing of the participant.

An eating event was defined by the following rules: (1) at least 50 kcal, and (2) a time gap to another food or meal of at least 15 minutes [23], while 10% of individual variability in both kcal and time were acceptable based on the individual nutritionist decision. Regarding the definition of a snack vs. meal and/or the size of meal, the following rules were set up based on the time of the day, nutrient density, and regularity. An eating event was defined as a “meal” if it has > 15.0 % of daily energy intake, and a snack if it comprised < 15.0% of daily energy intake [24]. Additionally, the time was considered, with main meals being between 08:00-10:00, 12:00 – 14:00, and 18:00 – 20:00, and snacks occuring outside of these “core eating times” [25]. Finally, consideration of nutrient density and regularity [26, 27] led to establishment of the following rules:

(1) 50-150 kcal = 5 minutes duration and separated by a 15-30 minute gap (small snack)

(2) 151-250 kcal = 10 minutes duration and separated by a 30-45 minute gap (big snack)

(3) 251-350 kcal = 15 minutes duration and separated by a 45-60 minute gap (small meal)

(4) 351-550 kcal = 20 minutes duration and separated by a 60-75 minute gap (normal meal)

(5) >550 kcal = 30 minutes duration and separated by a 75-90 minute gap (big meal)

For the definition of the first and the last meal as a breakfast and dinner, some deviations from these rules were made based on individual decisions. For example, if a person ate a bowl of oats with fruits every morning, which did not exceed 250 calories or 15.0 % of the individual’s energy intake, it was still considered breakfast due to its regularity and nutrient density. For foods reported after midnight, we used methods previously reported to assess evening eating. Caloric drinks that were not noted with any timing, but as a total consumed over the whole day were split among all meals of the day. The same procedure was used when meals were documented over a very long time that overlapped with other meals.

### 2.5. Chronotype and sleep time assessment

Chronotype and sleep timing were assessed by the Munich Chronotype Questionnaire [28]. Chronotypes were defined by the midpoint of sleep corrected for sleep-debt accumulated over the week (MSFsc) and classified as follows: early types: MSFsc ≤ 3:59; intermediate types: MSFsc = 4:00-4:59; late types: MSFsc ≥ 5:00 [29].

### 2.6. Statistical Analysis

Statistical analyses were carried out using SPSS 25.0 (IBM SPSS, Chicago, IL). Statistical significance was defined as p < 0.05. Data were expressed as mean (SD). Associations between meal timing and cardiometabolic parameters were examined using linear regression models. For this, not normally distributed variables were logarithmically transformed to bring their distributions closer to normal and used in subsequent analyses.

Heritability analysis of traits was based on the comparison of the similarity (correlation) of MZ twins with the similarity of DZ twins. That allowed to estimate the contribution of additive (A) genetic factors, as well as shared (C) and individual (E) environmental factors by structural equation models. For each variable, the full models (ACE) were estimated and tested against nested sub-models, where A and/or C components were fixed to zero, and the log-likelihood ratio test was used to compare the fit of the different models and sub-models to the observed data. Analysis was conducted by the Open-Mx package v. 2.21.11 for the R software v. 4.4.1. A, C, and E represent the proportion of variance explained by each component and sum up to 1 (or 100%). Results on heritability (A factor) were interpreted as follows [16]: < 0.20 for low heritability, 0.20 - 0.49 for moderate heritability, and ≥ 0.50 for high heritability.

## 3. Results

### 3.1. Clinical characteristics of participants

The eating pattern was assessed in 89 individuals (age: 31.9 (14.2) years, BMI: 22.8 (2.8) kg/m²) without diabetes (HbA1c: 5.01 (0.36) %) comprising 44 twin pairs, 32 monozygotic (MZ) and 12 dizygotic (DZ) pairs, with one individual without a pair in the MZ group. Anthropometric measures and metabolic characteristics are shown in the **Table 1**. The mean sleep onset took place at 23:16 (00:55) hr and sleep offset at 07:28 (01:09) hr, resulting in a total sleep duration of 7:52 (00:51) hours (**Table 1**). Among the participants, 38 showed an early, 24 - an intermediate, and 27 a late chronotype.

**Table 1.**
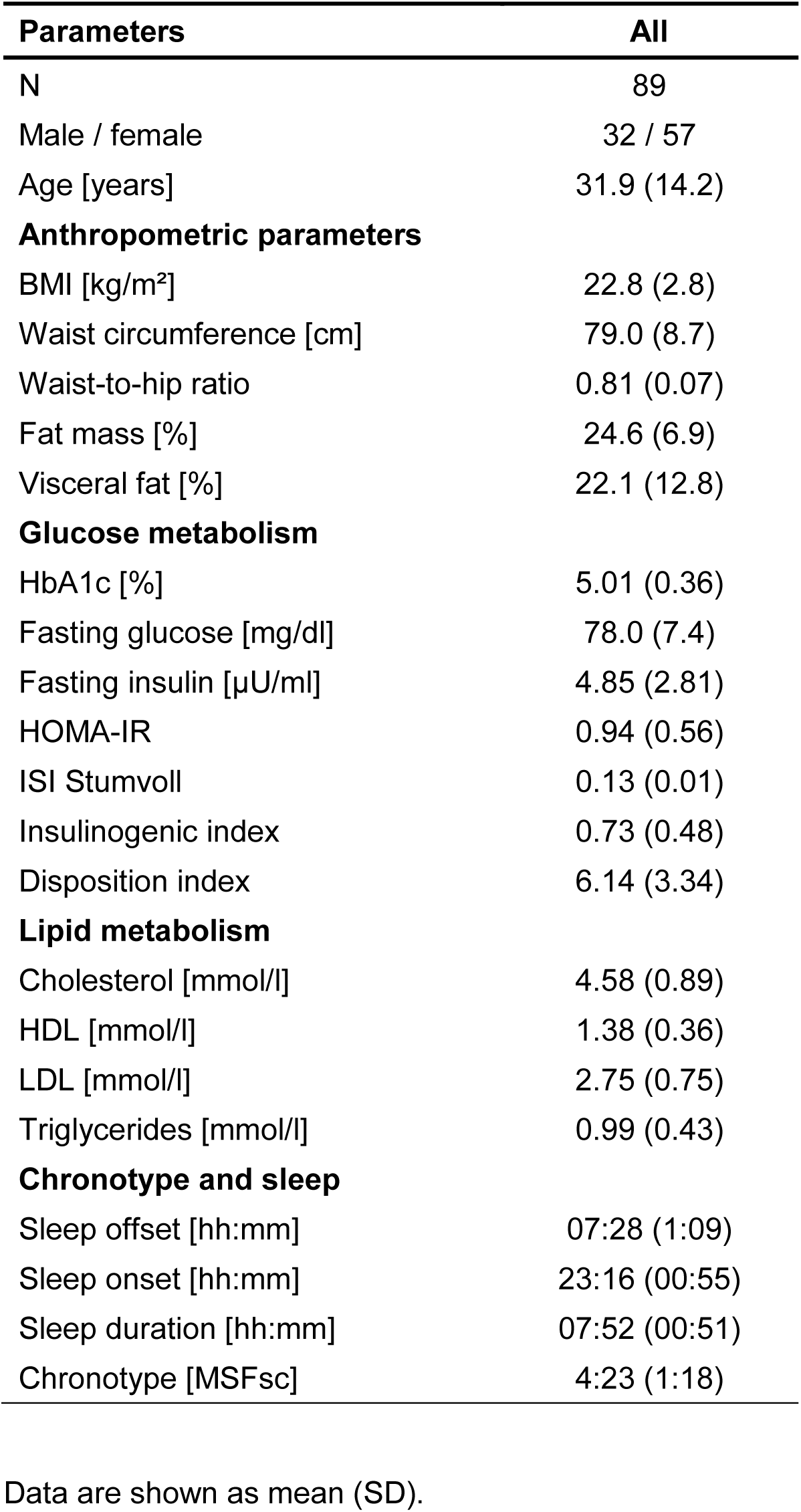
Clinical characteristics of study participants.

| **Parameters** | **All** |
| --- | --- |
| N | 89 |
| Male / female | 32 / 57 |
| Age [years] | 31.9 (14.2) |
| **Anthropometric parameters** |  |
| BMI [kg/m<sup>2</sup>] | 22.8 (2.8) |
| Waist circumference [cm] | 79.0 (8.7) |
| Waist-to-hip ratio | 0.81 (0.07) |
| Fat mass [%] | 24.6 (6.9) |
| Visceral fat [%] | 22.1 (12.8) |
| **Glucose metabolism** |  |
| HbA1c [%] | 5.01 (0.36) |
| Fasting glucose [mg/dl] | 78.0 (7.4) |
| Fasting insulin [μU/ml] | 4.85 (2.81) |
| HOMA-IR | 0.94 (0.56) |
| ISI Stumvoll | 0.13 (0.01) |
| Insulinogenic index | 0.73 (0.48) |
| Disposition index | 6.14 (3.34) |
| **Lipid metabolism** |  |
| Cholesterol [mmol/l] | 4.58 (0.89) |
| HDL [mmol/l] | 1.38 (0.36) |
| LDL [mmol/l] | 2.75 (0.75) |
| Triglycerides [mmol/l] | 0.99 (0.43) |
| **Chronotype and sleep** |  |
| Sleep offset [hh:mm] | 07:28 (1:09) |
| Sleep onset [hh:mm] | 23:16 (00:55) |
| Sleep duration [hh:mm] | 07:52 (00:51) |
| Chronotype [MSFsc] | 4:23 (1:18) |

Data are shown as mean (SD).

### 3.2. Meal timing pattern and interrrelations of its components

The daily distribution of eating events is shown in **Figure 1A-B**. The meal timing pattern was characterized by the assessment of the timing of meal consumption itself, calorie distribution across the day, and meal frequency (**Table 2**). Study subjects consumed 4.75 (1.15) meals per day, with the first eating event occurring at 09:15 (01:40) hr of the clock time and the last eating event at 20:12 (01:35) hr of the clock time, resulting in an eating window duration of 11:46 (02:07) hours (**Table 2**). Energy intake was 24.1 (8.7) % of daily calories in the first and 34.1 (10.7) % in the last meal, while the caloric midpoint of intake was estimated at 15:51 (01:39) hr of the clock time (**Table 2**).

**Table 2.** Meal timing pattern of study participants.

| Parameters | All |
| --- | --- |
| **Meal timing** |  |
| Eating window [hh:mm] | 11:46 (02:07) |
| Clock start of first eating event [hh:mm] | 09:15 (01:40) |
| Clock end of last eating event [hh:mm] | 20:12 (01:35) |
| Circadian start of first eating event [hh:mm]<sup>a</sup> | 04:52 (01:46) |
| Circadian end of last eating event [hh:mm]<sup>a</sup> | 08:10 (02:09) |
| **Daily calorie distribution** |  |
| Clock time of caloric midpoint [hh:mm] | 15:51 (01:39) |
| Circadian time of caloric midpoint [hh:mm]<sup>a</sup> | 12:32 (01:44) |
| Calories in first meal [%] | 24.1 (8.7) |
| Calories in last meal [%] | 34.1 (10.7) |
| **Meal frequency** |  |
| Number of meals | 4.75 (1.15) |

Data are shown as mean (SD).
<sup>a</sup> Circadian meal timing parameters were calculated as a time difference between the clock time of eating and MSFsc.

Multiple components of the meal timing pattern – especially clock start of first eating event, calories in the last meal and caloric midpoint - correlated with MSFsc and sleep offset and onset time (**Table S1**). In particular, later wake and sleeping time and correspondingly later chronotype were related to the later clock time of calorie midpoint and higher calorie intake during the last meal. This strong relation between the meal timing pattern and sleep-wake rhythm aligns with published data that an analysis of circadian meal timing in relation to the individual circadian phase is a more correct method compared to the clock time [30]. Therefore, in our study, we focussed on the circadian meal timing calculated as a time difference between the clock time of eating and MSFsc, a marker of individual’s chronotype (**Figure 1C)**. Circadian time of the first eating event was 19:07 (01:46) h, circadian time of the last eating event was 08:10 (02:09) h, and circadian time of the caloric midpoint was 12:32 (01:44) h.

Further, components of the meal timing pattern were strongly interrelated. There were positive associations of the duration of the eating window with total calorie intake and number of meals, as well as negative associations with a calorie intake during both first and last meal. This means that individuals with longer eating window consumed more calories in course of the day by eating more meals, but the individual meals, at least the first and last ones, were smaller (**Table S2**). There were also associations between the circadian time of the first eating event and all components of daily caloric distribution. In particular, the calorie intake during the first and last meal was less, and the circadian caloric midpoint was earlier, if the first eating event was earlier in relation to the MSFsc. In contrast, later circadian time of the last eating event positively correlated with a later circadian caloric midpoint (**Table S2**).

### 3.3. Association of circadian meal timing and glucose metabolism parameters

To assess the association of meal timing with parameters of glucose metabolism, i.e., insulin and glucose levels, insulin sensitivity and secretion indices in OGTT, a correlation analysis was conducted. Most associations were found for the circadian caloric midpoint, which showed a negative association with the fasting index of insulin resistance, HOMA-IR (β = - 0.358, p = 5.8 x 10^-4^), and a positive association with the OGTT-derived index of insulin sensitivity, ISI Stumvoll (β = 0.430, p = 2.6 x 10^-5^) (**Figure 2)**. Another OGTT-derived index of insulin sensitivity, the Matsuda index, was also positively associated with the circadian caloric midpoint (β = 0.263, p = 0.013) (**Table S3**). To account for potential confounders, the analysis model was adjusted for gender and age, as well as for energy intake and sleep duration. After adjustment, the associations with HOMA-IR (β = -0.276, p = 0.007) and ISI Stumvoll (β = 0.334, p = 2.9 x 10^-4^) remained significant **(Figure 3A-B)**, whereas association with the ISI Matsuda was lost in the last model (**Table S3**). The circadian caloric midpoint was also negatively associated with fasting insulin (β = -0.347, p = 8.7 x 10^-4^), AUC glucose (β = -0.250, p = 0.018), and AUC insulin (β = -0.256, p = 0.015) in OGTT, and positively related to the disposition index (β = 0.278, p = 0.008) (**Figure 2)**. These interrelations still persisted after the adjustment for gender and age, and for the fasting insulin - even after the additional adjustment for energy intake and sleep duration **(Figure 3C, Table S3)**.

**Figure 2.**
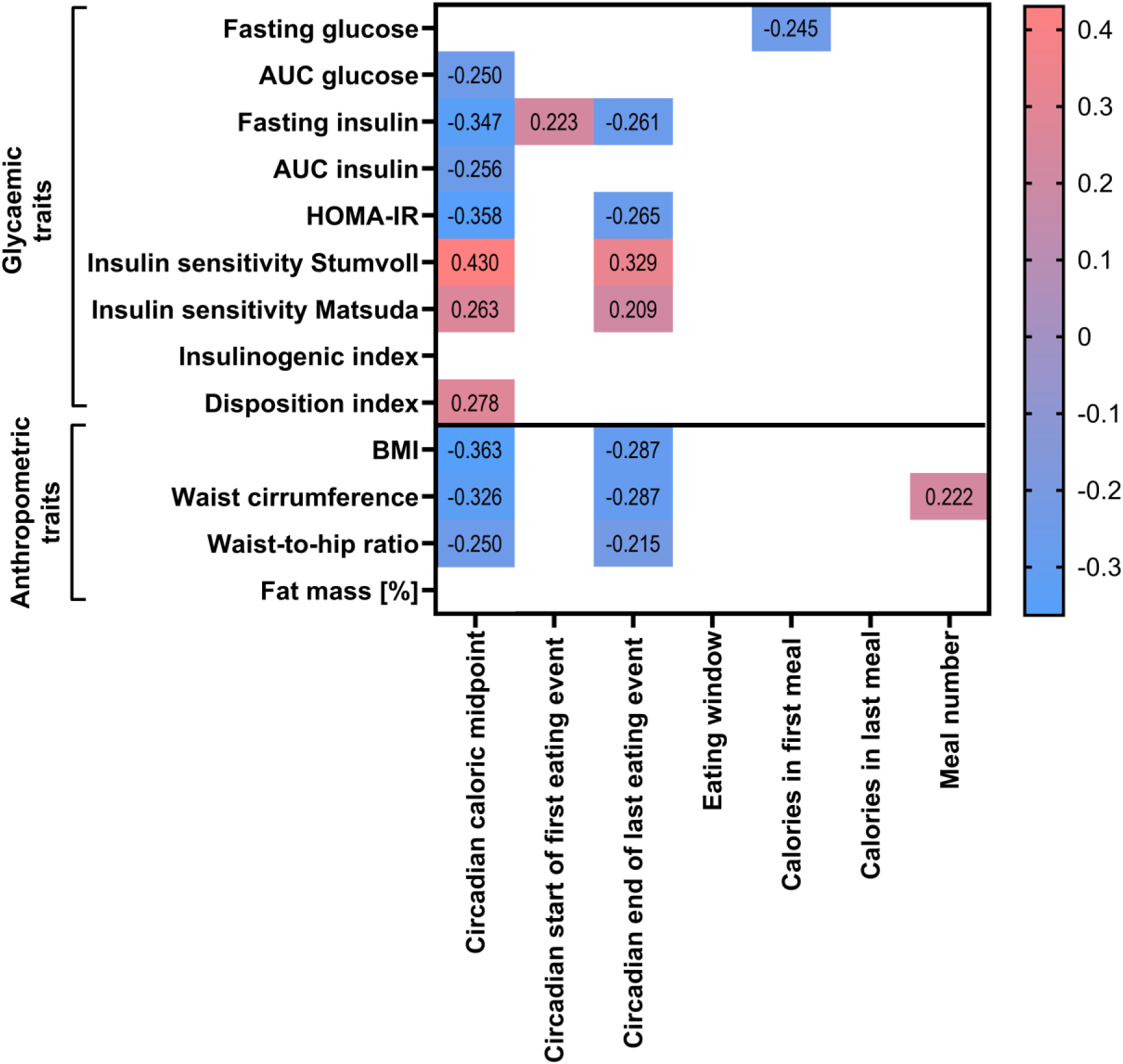
Association of meal timing and glycaemic parameters adjusted for potential cofounding factors. Heatmap represents beta-coefficient values in linear regression models that remained significant after the adjustment for gender, age, energy intake and sleep duration. Intensity of colour represents the strength of the correlation, whereas red indicates a positive and blue a negative correlation.

**Figure 3.**
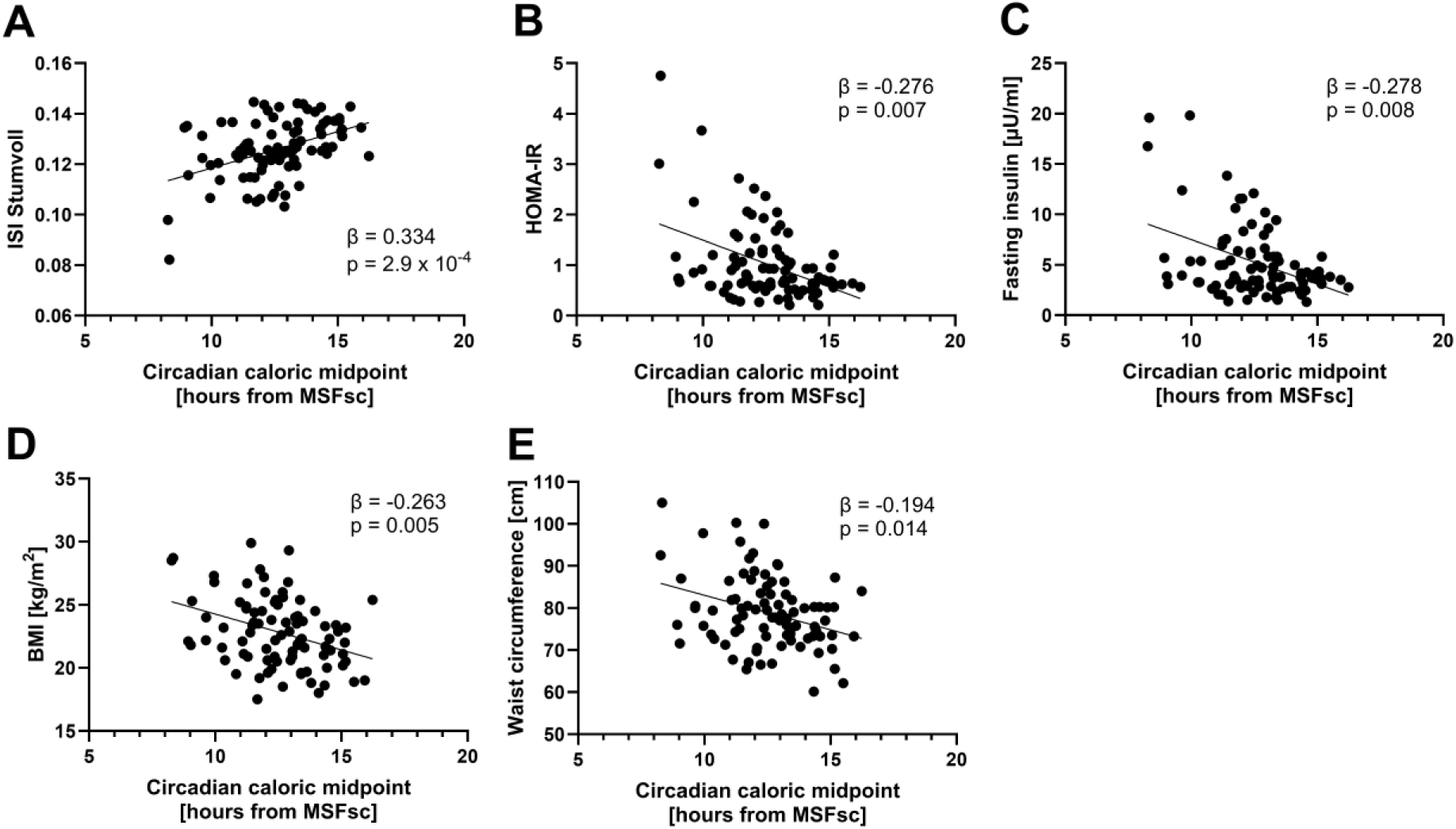
Associations of circadian caloric midpoint to insulin sensitivity, fasting insulin and anthropometric parameters. Associations with ISI Stumvoll (A), HOMA-IR (B), fasting insulin (C), BMI (D), and waist circumference (E) are shown. Beta-coefficient and p-values calculated using linear regression models adjusted for gender, age, energy intake and sleep duration are represented.

Similar to the circadian caloric midpoint, the circadian time of the last eating event showed positive associations with OGTT-derived insulin sensitivity indices (β = 0.329, p = 0.002 for ISI Stumvoll) and the disposition index (β = 0.210, p = 0.048), as well as negative associations with HOMA-IR (β = -0.265, p = 0.012) and fasting insulin (β = -0.261, p = 0.013) (**Figure 2)**, but no significance in adjusted models (**Table S4)**. Observed negative associations of the circadian start of the first eating event with fasting insulin and calorie intake during first meal with fasting glucose were also lost after adjustment for all cofounders (**Figure 2, Table S5)**.

Remarkably, the circadian time of the caloric midpoint and of the last eating event were negatively associated with BMI, waist circumference, and waist-to-hip ratio (**Figure 2**), consistent with previously published data [7, 30, 31]. The association with BMI (β = -0.363, p = 4.7x10^-4^) and waist circumference (β = -0.326, p = 0.002) remained significant after adjustment for gender, age, energy intake, and sleep duration (**Figure 3D-E, Table S3, S4)**. The association of daily meal number and waist circumference (β = 0.222, p = 0.036) was also observed in all adjusted models (**Figure 2, Table S5**). No associations with glycaemic or anthropometric traits were found for eating window duration and calorie intake during last meal.

### 3.4. Heritability of meal timing components

To estimate the contribution of genetic and environmental factors to the meal timing pattern, intrapair correlation coefficients for MZ and DZ were calculated (**Table S6**), and the A, C, and E parameters were assessed. MZ twins showed higher or similar intrapair correlations compared to DZ twins for meal timing parameters, except for eating window duration. Additionally, MZ twins showed higher intrapair correlations than DZ twins for all sleep timing parameters and chronotype. Accordingly, the variance of most meal and sleep timing parameters investigated in this study, is best explained by AE or ACE models (**Table S7**). In contrast, the duration of eating window showed a higher DZ correlation compared to MZ resulted in a better fit of the CE model (**Table S7**).

The estimation of heritability revealed higher heritability for the circadian start of the first eating event (59.2 %) than to the end of the last eating event (29.3 %), whereas the circadian caloric midpoint showed an intermediate value (34.5%). High or moderate heritability was shown both for the percent of energy in the first (43.9%) and last meal (59.7 %), as well as in meal number (54.4 %) (**Figure 4**). Remarkably, most sleep parameters strongly related to the meal timing pattern also showed high (52.7 % for sleep onset, 57.8% for sleep duration, and 57.3 % for MSFsc) or moderate (41.3 % for sleep offset) heritability (**Figure 4**).

**Figure 4.**
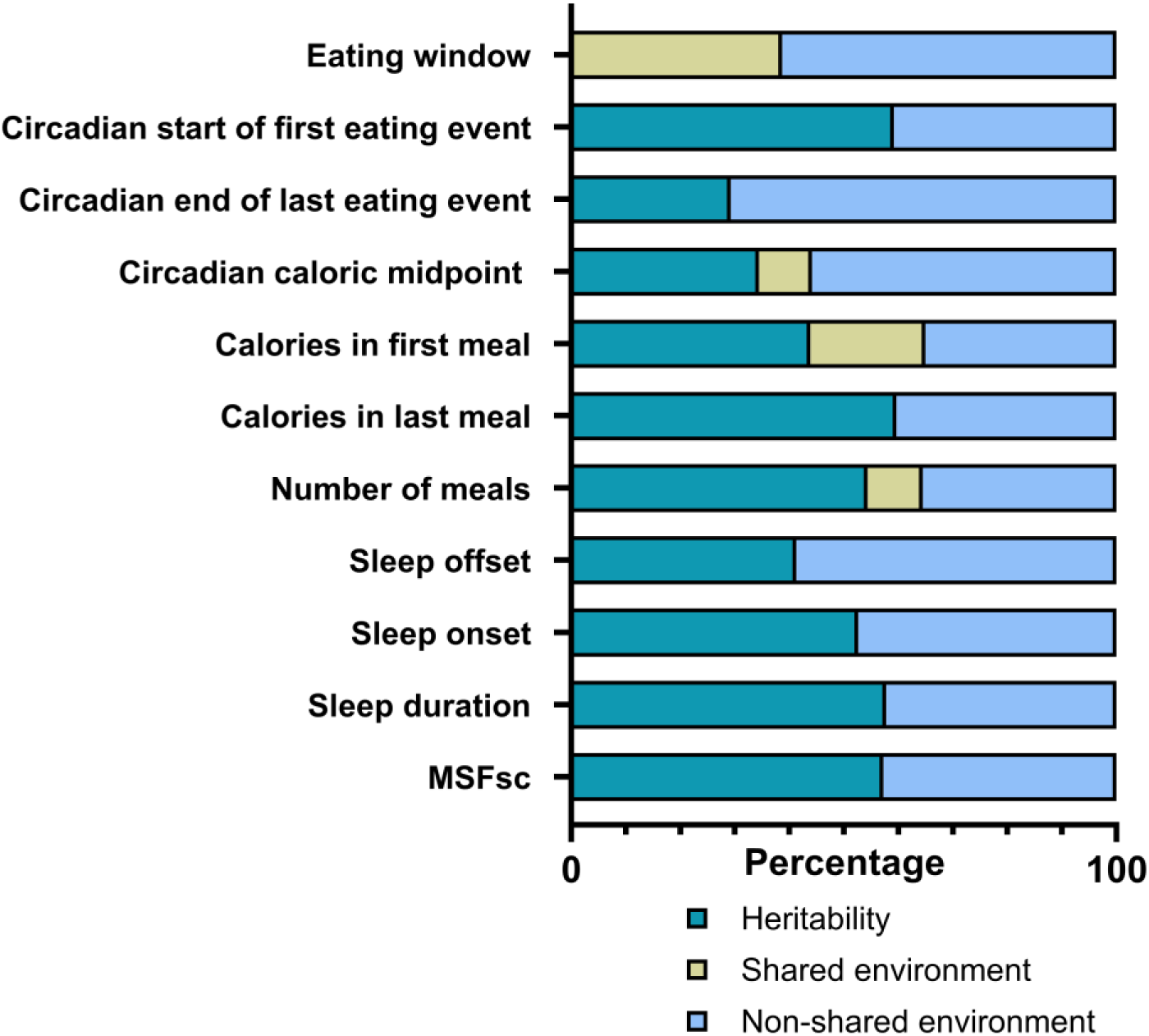
Heritability of meal timing and sleep parameters. The percentage of heritability is represented in petrol (additive genetic factors, A), and environmental factor contribution is shown in khaki (shared environmental factors, C) and blue (non-shared environmental factors, E).

## 4. Discussion

In this study, we demonstrated for the first time robust associations of the circadian meal timing with insulin sensitivity representing one of key players in the development of type 2 diabetes [32]. We also expanded current knowledge on the heritability of meal timing patterns, observing the essential genetic components contributing to the individual timing of eating throughout of the day.

The highlight of this study is the detailed characterization of individual meal timing patterns by assessment of timing of meal consumption itself, meal frequency, and the diurnal distribution of energy intake. Notably, recent research has shown that using clock hours to define meal timing may be misleading because of individual differences in circadian phase. Indeed, specific dinner timing can be classified as late for early chronotype and early for the late chronotype [33] and might therefore have a different impact on metabolic state and/or risks. Therefore, the assessment of circadian meal timing in relation to the individual circadian phase represents a state-of-art personalized approach in chrononutritional research [30, 33, 34], and should be preferred if chronotype, dim light melatonin onset (DLMO), or sleep timing data are available. In present study, circadian timing of meals was determined as the time difference in relation to the midpoint of sleep (MSFsc) [28] and showed multiple associations with parameters of glycose metabolism.

Most associations with glycaemic parameters were found for the circadian caloric midpoint, which was also negatively correlated with BMI and waist circumference, even after adjusting for gender and age as well as for energy intake and sleep duration. The circadian timing of the last eating event showed similar associations. This is in agreement with previous research revealed that late caloric midpoint in relation to DLMO or sleep onset is associated with obesity-related traits such as BMI and body fat [7, 30, 31]. Remarkably, late eating can also result in less effective weight loss on a hypocaloric diet compared to eating early at the day [35]. Late dinner timing was also shown to be associated with impaired glucose tolerance, which might be partly explained by the concurrence of food intake and elevated concentrations of endogenous melatonin [36]. Researchers have shown that evening intake of large, high glycaemic index (GI) meals [37, 38] as well as low-GI meals [39], can negatively affect postprandial glucose profiles. These profiles can even resemble the response of (pre)diabetic individuals, even if the participant was healthy [40], supporting the assumption that late food intake may play a role in increasing the risk of type 2 diabetes. Taking together, the caloric midpoint is obviously a key meal timing parameter strongly contributing to the obesity and diabetes risk.

The important novelty of our study is the analysis of OGTT-derived glucose metabolism parameters in relation to meal timing. Specifically, we assessed both the insulin sensitivity and insulin secretion indices, as well as a disposition index reflecting a beta-cell function. Our study for the first time described the negative associations of circadian caloric midpoint with fasting insulin and HOMA-IR, which are simple fasting measures of insulin resistance, as well as a strong positive association with an OGTT-derived Stumvoll index of insulin sensitivity. All these associations remained significant after adjustment for potential cofounders. Previously, the association of higher HOMA-IR with late eating was shown in the NHANES cohort [41] and a relatively small cross-sectional study [42]. However, the association of caloric midpoint with insulin sensitivity after a glucose challenge (i.e., in OGTT) had not been demonstrated until now. The additional assessment of other ISI proposed by Matsuda confirmed our finding. This novel finding underscores the essential role of individual diurnal calorie distribution not only to obesity, but also to diabetes risk.

Further, both glucose and insulin levels in OGTT were negatively interrelated with circadian caloric midpoint, suggesting better glucose tolerance in individuals who consume most calories earlier in the day. Next, the disposition index which is used as a measure of beta cell capability to compensate for insulin resistance and predicts the conversion of insulin resistance to diabetes [43], showed a positive association with circadian timing of the caloric midpoint and of the last eating event. This suggest that consuming most calories early in the day may be protective against diabetes. In our results, the change of the disposition index clearly reflects the insulin sensitivity described above, while insulin secretion was less or not at all linked to circadian meal timing. Finally, the negative association between energy intake during the first meal and fasting glucose observed in our study aligns with data on beneficial effects of a high-calorie breakfast on glycaemic control in individuals with type 2 diabetes [44, 45], underscoring the essential role of diurnal calorie distribution in metabolic regulation. Notably, all associations, mentioned in this paragraph, lost the significance after the adjustment for gender, sex, energy intake, and sleep duration, indicating that the circadian calorie midpoint with insulin sensitivity is independent of these cofounding factors.

Another robust association found in our study is the interrelation between the daily number of meals and waist circumference, which remained significant after adjustment for total energy intake and might be partly explained by reduced fasting periods between meals. Regular fasting periods provide metabolic flexibility and a number of other physiological benefits such as reduced inflammation, improved circadian rhythmicity, increased autophagy, and modulation of the gut microbiota [46] and may therefore protect against diabetes.

Surprisingly, our study revealed no correlations between the eating window duration and any glycaemic or obesity-related anthropometric traits, despite expectations based on interventional time-restricted eating (TRE) studies. Indeed, the link between the daily eating duration and body weight has been shown in TRE trials often accompanied by the spontaneous reductions in calorie intake [47], but not in observational or prospective human cohorts. The role of eating duration therefore needs to be further investigated in large cohort studies.

The next importance of our study is that it expands the knowledge that the meal timing pattern is influenced by genetic factors. Emerging data have demonstrated the heritability of food intake parameters, ranging from overall energy intake [48] to more subtle variables known as the microstructure of food intake, including frequency, composition, and meal timing, independent of overall intake [49]. Twin studies have shown that genetics influence various diet-related phenotypes, such as energy and macronutrients intakes, dietary patterns, and the intake of specific food items [50]. Additionally, specific physiological variables, such as stomach filling before and after eating [51], and psychological variables, such as the subjective rating of hunger and palatability of food, also show genetic influences [52].

However, data on the heritability of meal timing are currently very limited. Our study revealed high heritability (over 50 %) was shown for most parameters of the meal timing pattern, including circadian timing of the first eating event, the percentage of energy consumed in the last meal, and meal number. Remarkably, the circadian timing of the last eating event and of the caloric midpoint demonstrated lower genetic influences. Meal timing components were strongly related to individual sleep-wake timing and chronotype, which also showed a marked genetic impact. This suggests that these traits may share a similar genetic architecture. Given this genetic contribution, modifying individual meal timing patterns might be challenging due to the high heritability and its relation to sleep-wake rhythms. However, shifting of the caloric midpoint and the last meal timing to earlier in the day might be more feasible compared to the start of eating, due to the lower heritability.

Our data partly align with the findings of [16], which reported high heritability (64 %) for the caloric midpoint, 56% for breakfast, and 38 % for lunch. Together with their findings for the heritability of sleeping behaviour, with 55 % for wake time and 38 % for bedtime, the authors conclude that heritability estimates are higher for behaviour earlier in the day. This contrasts with other recent research [17], which did not show a significant difference in the genetic contribution to morning and evening behaviours, reporting heritability of 24% for morning, 18% for afternoon, and 22% for evening periods. The discrepancies in the estimation of the genetic component could be explained by differences in the study populations. The Spanish study population in [16] was older and overweight, whereas the NUGAT cohort consisted of younger and lean individuals.

Specific genetic factors contributing to meal timing include genetic polymorphisms. As an example, obese participants carrying the minor G allele of the CLOCK gene rs4580704 variant had their lunch significantly later than participants carrying major allele [35]. The obesity-related PLIN1 gene polymorphism rs1052700 seems to influence the interaction between meal timing and weight loss: AA carriers who ate lunch late lost less weight than those who ate early, whereas the time of eating did not influence weight loss among TT carriers [53]. Moreover, genetic influences have been suggested for the night eating syndrome and sleep-related eating disorders, which are characterized by a preference for late-night eating [54]. Contrary to these findings, the first genome-wide association study for breakfast skipping (TwinUK) including 2006 participants did not find any variants that surpassed the genome-wide significance threshold [55]. Therefore, it is still tempting to elucidate the genes associated with mistimed food intake, as these may help to identify individuals who are at higher risk of mistimed eating or, conversely, resistant to timing dietary interventions.

Finally, as confirmed by our results, beside of genetic contribution, a number of environmental factors, including cultural aspects, determine meal timing pattern [14, 15]. For instance, the caloric midpoint assessed in the German NUGAT cohort (15:51 hr) slightly differs from those shown by Lopez-Minguez et al. (15:20 hr) [16] and Teixeira et al. (15:00 hr) [56], which might be explained by cultural or regional differences. Further studies are needed to differentiate modifiable and non-modifiable (e.g., genetic) factors that determine meal timing pattern and can be effectively targeted in the dietary intervention studies.

### 4.1. Limitations/strengths

The marked strength of our study is a detailed metabolic, circadian and dietary habit phenotyping in a NUGAT cohort which allowed to both assess the association of meal timing with glycaemic traits and explore the contribution of genetic factors to the meal timing architecture. The further strength of our study, as detailed above, is the analysis of multiple components of the meal pattern considered in relation to the individual circadian phase. The final strength is the inclusion of both genders enhancing the generalizability of our findings. This contrasts with the cohort of Lopez-Minguez et al. [16] limited to adult females with a slightly increased BMI.

Some limitations have to be discussed when interpreting the results of this work. Food as well as sleep timing were self-reported and are therefore prone to measurement errors as over- and underreporting. However, the reporting behaviour of our cohort remained largely the same, and other data was collected by a study team under clinical supervision. Finally, the sample size of 92 can be seen as a further limitation when comparing this to similar studies, where sample size went up to 265 for [17]. Notably, the sample size of Lopez-Minguez et al. [16] with 106 participants did not exceed ours by far.

## 5. Conclusions

Our study revealed that the circadian meal timing is associated with insulin sensitivity and shows significant genetic influences. Multiple associations between the circadian caloric midpoint and glycaemic parameters, as well as BMI, persisted even after adjustment for cofounding factors, highlighting the importance of aligning main calorie intake to an earlier circadian time that matches individual circadian rhythms. Modifying calorie distribution in this way might protect against diabetes, although this could be challenging due to the high heritability of meal timing components.

## Data Availability

All data produced in the present study are available upon reasonable request to the authors

## Acknowledgements

We thank all study participants for their cooperation. We gratefully acknowledge the technical assistance of Andreas Wagner, Melanie Hannemann and Anja Henkel. We also thank Elwira Gliwska for her contribution the algorithm development for the analysis of meal timing.

## Author contributions

Study concept and design: MK, AFHP, OP-R. Acquisition of data: JV, BP, SH, A-CO. Analysis and interpretation of data: JV, BP, AB, OP-R. Drafting, editing, and revision of the manuscript: JV, BP, AFHP, OP-R. All authors read and approved the final manuscript.

## Funding

This work was supported by the German Research Foundation (DFG RA 3340/4-1 to OP-R, project number 530918029), by the European Association for the Study of Diabetes (Morgagni Prize 2020 to OP-R), and by the German Federal Ministry of Education and Research (BMBF NUGAT 0 315 424 to AFHP).

## Declaration of competing interest

The authors have nothing to disclose.

## Abbreviations

AUC: Area under the curve
BMI: Body mass index
CLOCK: Circadian locomotor output cycles kaput gene
DZ: Dizygotic
ELISA: Enzyme linked immunosorbent assay
HbA1c: Haemoglobin A1c
HDL: High-density lipoprotein
HOMA-IR: Homeostatic Model Assessment for Insulin Resistance
ISI: Insulin sensitivity index
LDL: Low-density lipoprotein
MSFsc: Mid-sleep on free days, sleep corrected
MCTQ: Munich Chronotype Questionnaire
MZ: Monozygotic
NUGAT: NUtriGenomics Analysis in Twins
OGTT: Oral glucose tolerance test
PLIN1: Perilipin-1
SD: Standard deviation
SNP: Single nucleotide polymorphism
T2D: Type 2 diabetes

---

**Figure S1.**
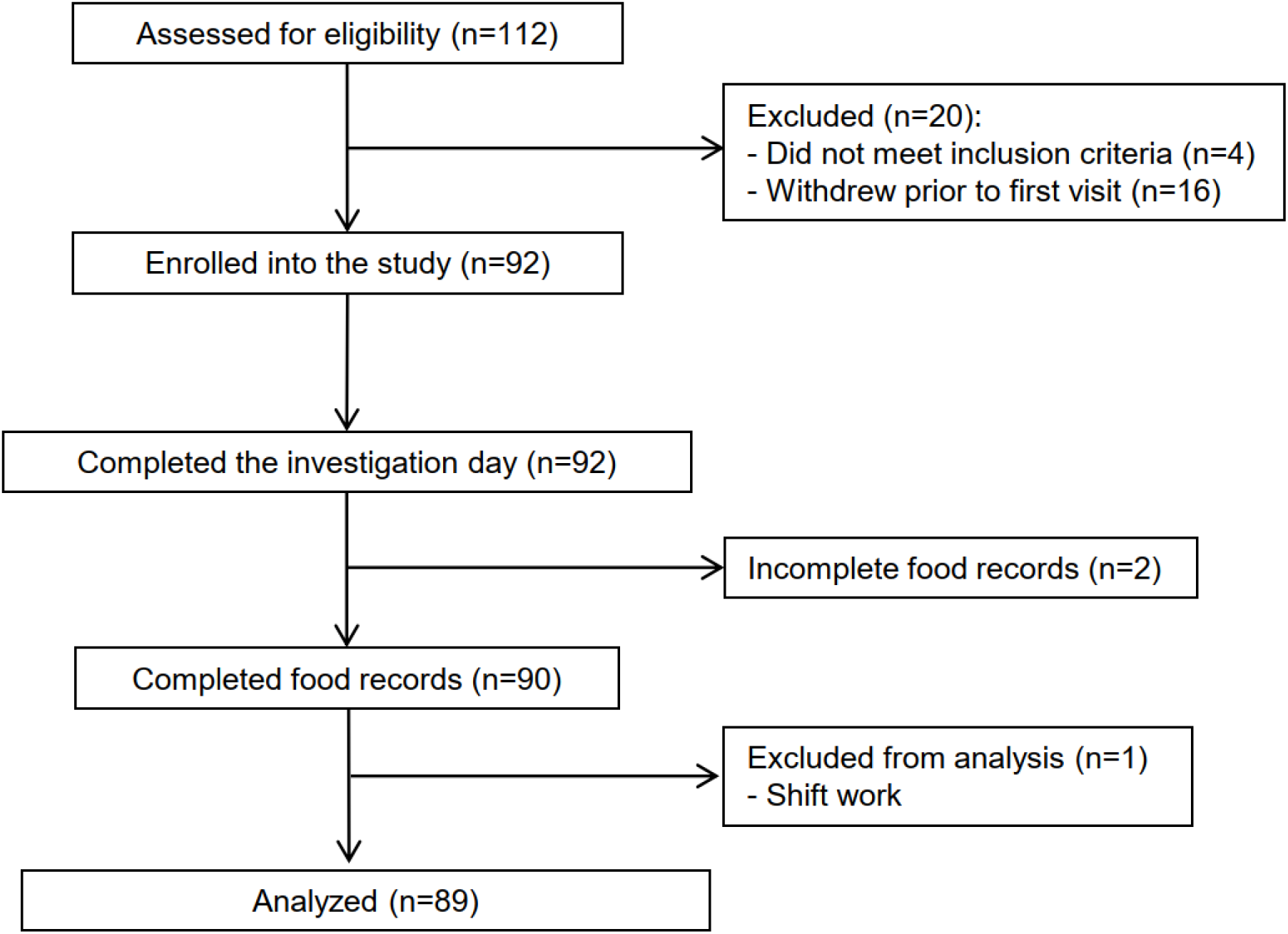
Study flow chart.

**Table S1.** Correlation analysis of meal and sleep timing parameters.

|  | Sleep offset | Sleep onset | Sleep duration | MSFsc |
| --- | --- | --- | --- | --- |
| **Meal timing** |  |  |  |  |
| Eating window | -0.361** |  |  |  |
| Clock start of first eating event | 0.408*** | 0.349*** |  | 0.335** |
| Clock end of last eating event |  |  |  |  |
| **Calorie distribution** |  |  |  |  |
| Total calorie intake |  |  |  |  |
| Calories in first meal |  |  |  |  |
| Calories in last meal | 0.326** | 0.298** |  | 0.280** |
| Caloric midpoint | 0.314** | 0.351*** |  | 0.325** |
| **Meal frequency** |  |  |  |  |
| Number of meals | -0.300** |  |  |  |

Only significant correlation coefficients are shown. \* if $p < 0.05$ . \*\* if $p < 0.01$ . and \*\*\* if $p < 0.001$ .

**Table S2.**
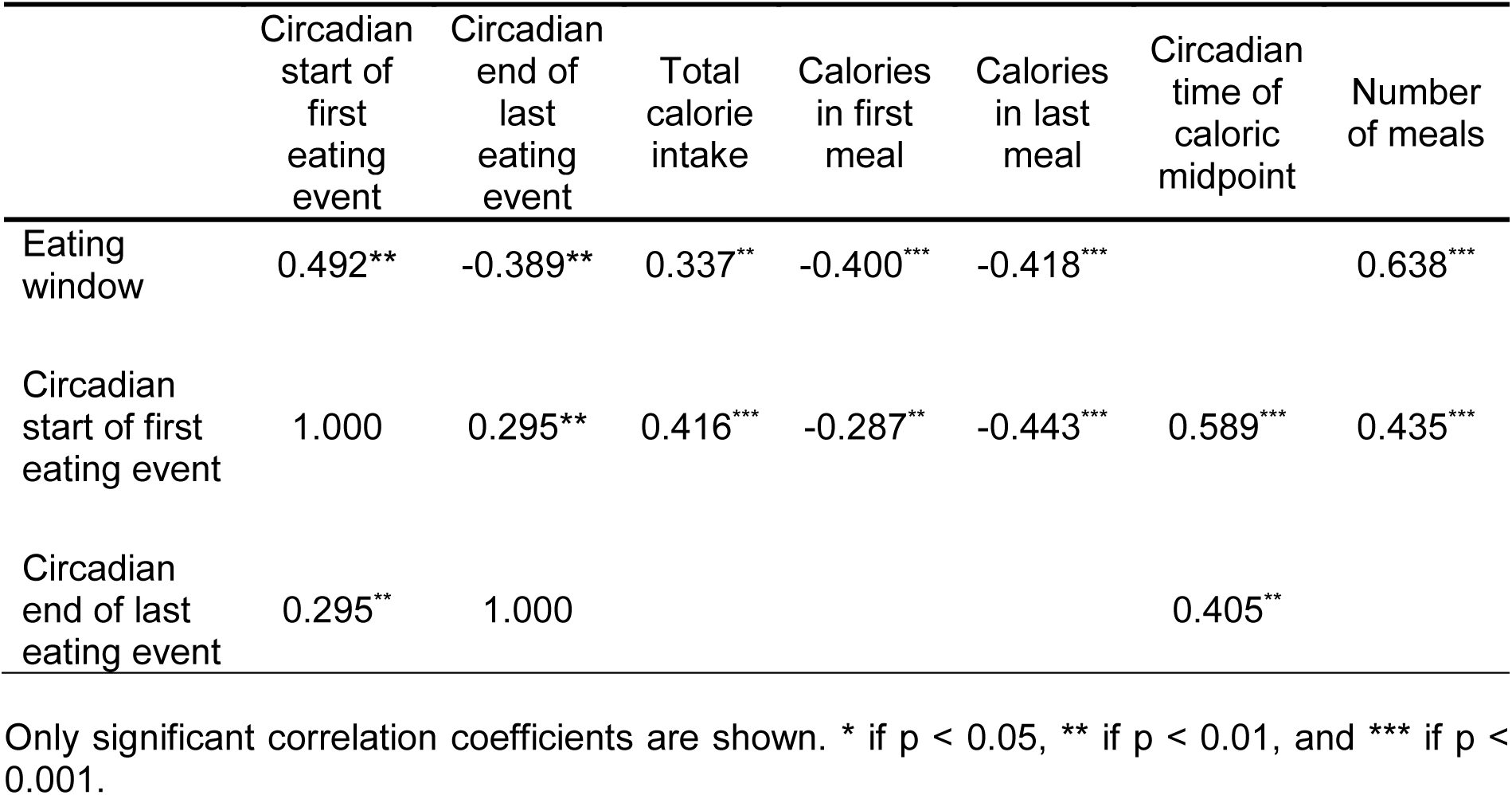
Correlation analysis of meal timing parameters and daily calorie distribution.

**Table S3.** Associations of circadian caloric midpoint with glycaemic and anthropometric traits.

|  | Model 1 |  |  | Model 2 |  |  | Model 3 |  |  |
| --- | --- | --- | --- | --- | --- | --- | --- | --- | --- |
| | $\beta$ | p | 95 % CI | $\beta$ | p | 95 % CI | $\beta$ | p | 95 % CI |
| **Glycaemic traits** |  |  |  |  |  |  |  |  |  |
| Fasting glucose | -0.188 | 0.077 | -0.099, 0.005 | -0.086 | 0.360 | -0.067, 0.025 | -0.090 | 0.352 | -0.070, 0.025 |
| AUC glucose | -0.250 | **0.018** | -0.037, -0.004 | -0.202 | **0.048** | -0.032, -0.00016 | -0.205 | 0.051 | -0.033, 0.00004 |
| Fasting insulin | -0.347 | **0.001** | -0.185, -0.050 | -0.300 | **0.004** | -0.169, -0.034 | -0.278 | **0.008** | -0.163, -0.025 |
| AUC insulin | -0.256 | **0.015** | -0.146, -0.016 | -0.239 | **0.027** | -0.143, -0.009 | -0.202 | 0.062 | -0.131, 0.003 |
| HOMA-IR | -0.358 | **0.001** | -0.199, -0.057 | -0.297 | **0.003** | -0.176, -0.036 | -0.276 | **0.007** | -0.170, -0.028 |
| ISI Stumvoll | 0.430 | **2.6x10<sup>-5</sup>** | 0.002, 0.004 | 0.346 | **1.7x10<sup>-4</sup>** | 0.001, 0.004 | 0.334 | **2.9x10<sup>-4</sup>** | 0.001, 0.003 |
| ISI Matsuda | 0.263 | **0.013** | 0.157, 1.283 | 0.230 | **0.032** | 0.056, 1.202 | 0.204 | 0.061 | -0.026, 1.141 |
| Insulinogenic index | -0.050 | 0.639 | -0.093, 0.058 | -0.041 | 0.709 | -0.092, 0.063 | -0.005 | 0.962 | -0.079, 0.076 |
| Disposition index | 0.211 | **0.048** | 0.001, 0.160 | 0.163 | 0.121 | -0.017, 0.141 | 0.184 | 0.087 | -0.010, 0.151 |
| **Anthropometric traits** |  |  |  |  |  |  |  |  |  |
| BMI | -0.363 | **4.7x10<sup>-4</sup>** | -0.881, -0.258 | -0.269 | **0.004** | -0.702, -0.142 | -0.263 | **0.005** | -0.693, -0.131 |
| Waist circumference | -0.326 | **0.002** | -0.032, -0.008 | -0.198 | **0.012** | -0.022, -0.003 | -0.194 | **0.014** | -0.021, -0.002 |
| Waist-to-hip ratio | -0.250 | **0.018** | -0.259, -0.025 | -0.127 | 0.123 | -0.165, 0.020 | -0.138 | 0.099 | -0.172, 0.015 |
| Fat mass | -0.064 | 0.549 | -1.101, 0.589 | -0.103 | 0.152 | -0.970, 0.153 | -0.065 | 0.346 | -0.799, 0.283 |

Linear regression analyses were performed to estimate the strength of association between variables. Model 1 is without any covariate adjustment. model 2 is adjusted for gender and age, and model 3 for total energy intake and sleep duration. $\beta$ are the standardized coefficients; 95% CI. P-values are marked with \* if $p < 0.05$ and \*\* if $p < 0.01$ .

**Table S4.** Associations of circadian time of the last eating event with glycaemic and anthropometric traits.

|  | Model 1 |  |  | Model 2 |  |  | Model 3 |  |  |
| --- | --- | --- | --- | --- | --- | --- | --- | --- | --- |
| | $\beta$ | p | 95 % CI | $\beta$ | p | 95 % CI | $\beta$ | p | 95 % CI |
| **Glycaemic traits** |  |  |  |  |  |  |  |  |  |
| Fasting glucose | -0.119 | 0.268 | -0.067, 0.019 | 0.029 | 0.765 | -0.034, 0.045 | 0.028 | 0.780 | -0.034, 0.046 |
| AUC glucose | -0.206 | 0.052 | -0.027, 0.000 | -0.087 | 0.420 | -0.020, 0.008 | -0.088 | 0.424 | -0.020, 0.008 |
| Fasting insulin | -0.261 | **0.013** | -0.128, -0.015 | -0.198 | 0.073 | -0.114, 0.005 | -0.188 | 0.088 | -0.111, 0.008 |
| AUC insulin | -0.189 | 0.076 | -0.102, 0.005 | -0.164 | 0.153 | -0.100, 0.016 | -0.153 | 0.174 | -0.096, 0.018 |
| HOMA-IR | -0.265 | **0.012** | -0.137, -0.017 | -0.181 | 0.093 | -0.114, 0.009 | -0.172 | 0.111 | -0.111, 0.012 |
| ISI Stumvoll | 0.329 | **0.002** | 0.001, 0.003 | 0.178 | 0.074 | 0.000, 0.002 | 0.176 | 0.074 | 0.000, 0.002 |
| ISI Matsuda | 0.209 | **0.049** | 0.002, 0.930 | 0.160 | 0.158 | -0.141, 0.853 | 0.151 | 0.182 | -0.160, 0.832 |
| Insulinogenic index | -0.045 | 0.673 | -0.074, 0.048 | -0.059 | 0.611 | -0.083, 0.049 | -0.050 | 0.660 | -0.080, 0.051 |
| Disposition index | 0.210 | **0.048** | 0.001, 0.130 | 0.116 | 0.296 | -0.032, 0.104 | 0.120 | 0.282 | -0.031, 0.106 |
| **Anthropometric traits** |  |  |  |  |  |  |  |  |  |
| BMI | -0.287 | **0.006** | -0.625, -0.105 | -0.149 | 0.132 | -0.438, 0.058 | -0.150 | 0.124 | -0.436, 0.054 |
| Waist circumference | -0.287 | **0.006** | -0.024, -0.004 | -0.129 | 0.123 | -0.015, 0.002 | -0.130 | 0.117 | -0.015, 0.002 |
| Waist-to-hip ratio | -0.215 | **0.043** | -0.195, -0.003 | 0.409 | -0.072 | -0.113, 0.046 | 0.374 | -0.078 | -0.115, 0.044 |
| Fat mass | -0.128 | 0.231 | -1.097, 0.268 | -0.039 | 0.609 | -0.610, 0.359 | -0.029 | 0.685 | -0.551, 0.364 |

Linear regression analyses were performed to estimate the strength of association between variables. Model 1 is without any covariate adjustment. model 2 is adjusted for gender and age, and model 3 for total energy intake and sleep duration. $\beta$ are the standardized coefficients; 95% CI. P-values are marked with \* if $p < 0.05$ and \*\* if $p < 0.01$ .

**Table S5.** Associations of other meal timing parameters with glycaemic and anthropometric traits.

|  | Model 1 |  |  | Model 2 |  |  | Model 3 |  |  |
| --- | --- | --- | --- | --- | --- | --- | --- | --- | --- |
| | $\beta$ | p | 95 % CI | $\beta$ | p | 95 % CI | $\beta$ | p | 95 % CI |
| **Circadian time of the first eating event** |  |  |  |  |  |  |  |  |  |
| Fasting insulin | -0.223 | **0.036** | 0.005, 0.143 | -0.252 | **0.014** | 0.017, 0.150 | -0.218 | 0.057 | -0.002, 0.147 |
| **Calories in the first meal** |  |  |  |  |  |  |  |  |  |
| Fasting glucose | -0.245 | **0.021** | -0.683, -0.058 | -0.157 | 0.091 | -0.514, 0.039 | -0.175 | 0.085 | -0.514, 0.039 |
| **Meal number** |  |  |  |  |  |  |  |  |  |
| Waist circumference | 0.222 | **0.036** | 0.006, 0.176 | 0.165 | **0.040** | 0.003, 0.133 | 0.244 | **0.008** | 0.003, 0.133 |

Linear regression analyses were performed to estimate the strength of association between variables. Model 1 is without any covariate adjustment. model 2 is adjusted for gender and age, and model 3 for total energy intake and sleep duration. Only parameters showing significant associations in model 1 are shown. $\beta$ are the standardized coefficients; 95% CI. P-values are marked with \* if $p < 0.05$ and \*\* if $p < 0.01$ .

**Table S6.** Twin intra-pair correlations of meal timing and sleep parameters.

|  | **rMZ** | **rDZ** |
| --- | --- | --- |
| **Meal timing** |  |  |
| Eating window | 0.043 | 0.639* |
| Circadian start of first eating event | 0.458** | 0.358 |
| Circadian end of last eating event | 0.273 | 0.146 |
| **Daily calorie distribution** |  |  |
| Circadian time of caloric midpoint | 0.464** | 0.207 |
| Calories in first meal | 0.585*** | 0.572 |
| Calories in last meal | 0.542* | 0.325 |
| **Meal frequency** |  |  |
| Number of meals | 0.408* | 0.604* |
| **Chronotype and sleep** |  |  |
| Sleep offset | 0.426* | -0.085 |
| Sleep onset | 0.527** | -0.204 |
| Sleep duration | 0.547*** | 0.328 |
| Chronotype [MSFsc] | 0.582*** | -0.029 |

Intra-pair correlations are shown for monozygotic (rMZ) and dizygotic (rDZ) twins. \* $p < 0.05$ , \*\* $p < 0.01$ ; \*\*\* $p < 0.001$ .

**Table S7.**
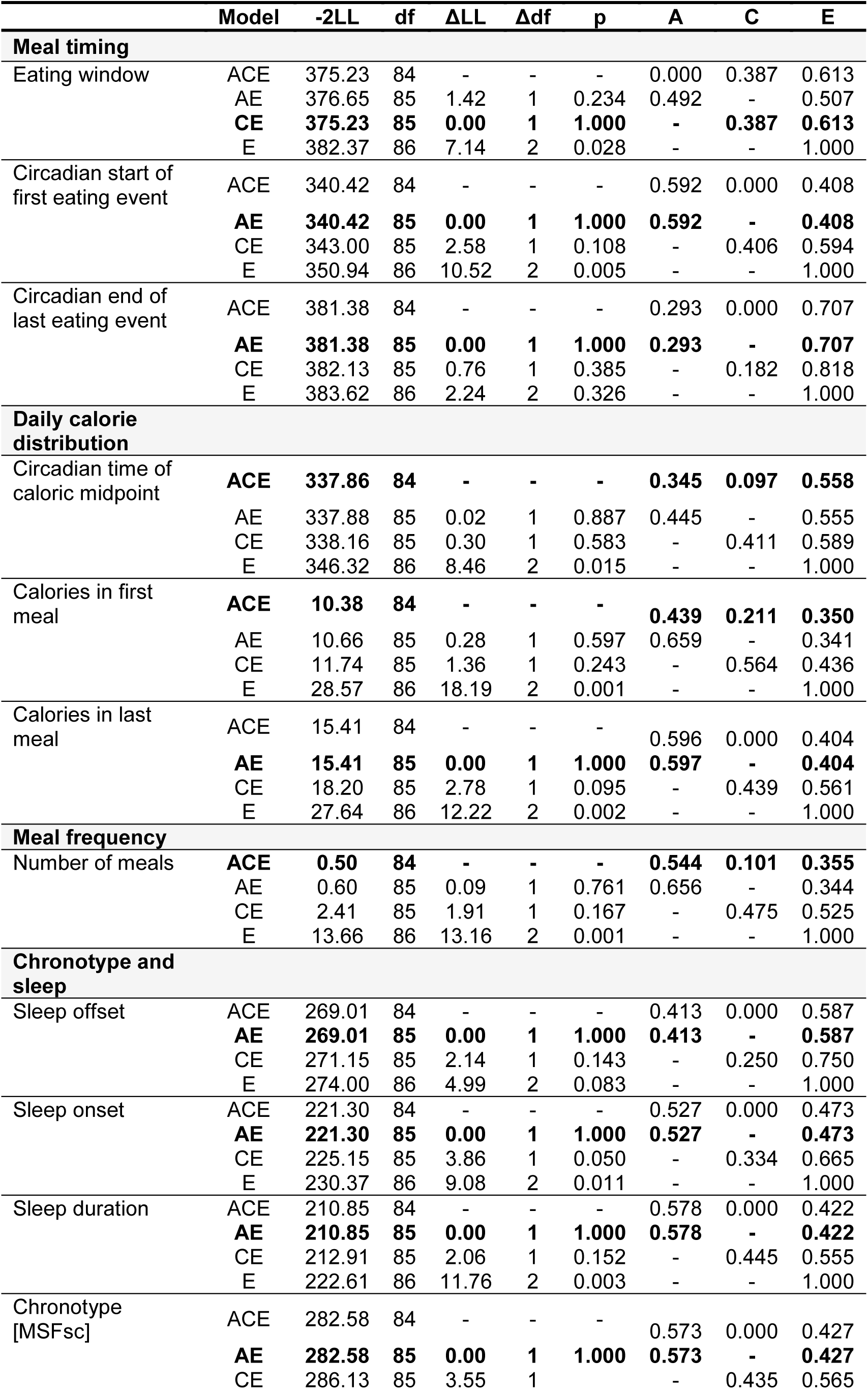
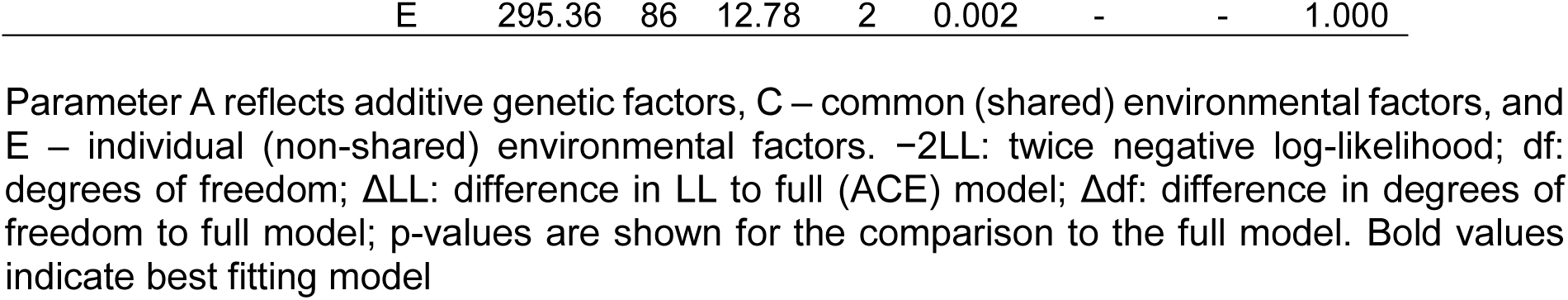
Model fitting and proportions of variance explained by genetic and environmental influences.

## Notes

### Competing Interest Statement

The authors have declared no competing interest.

### Clinical Trial

NCT01631123

### Author Declarations

Ethics committee of the Charite-Universitaetsmedizin Berlin gave ethical approval for this work

### Summary of Updates

Graphical abstract was revised; other parts of the manuscript were not changed

